# Studying the context of psychoses to improve outcomes in Ethiopia (SCOPE): protocol paper

**DOI:** 10.1101/2023.10.10.23296817

**Authors:** Charlotte Hanlon, Tessa Roberts, Eleni Misganaw, Ashok Malla, Alex Cohen, Teshome Shibre, Wubalem Fekadu, Solomon Teferra, Derege Kebede, Adiyam Mulushoa, Zerihun Girma, Mekonnen Tsehay, Dessalegn Kiross, Crick Lund, Abebaw Fekadu, Craig Morgan, Atalay Alem

## Abstract

**Background:** Global evidence on psychosis is dominated by studies conducted in Western, high-income countries. The objectives of the Study of Context Of Psychoses to improve outcomes in Ethiopia (SCOPE) are (1) to generate rigorous evidence of psychosis experience, epidemiology and impacts in Ethiopia that will illuminate aetiological understanding and (2) inform development and testing of interventions for earlier identification and improved first contact care that are scalable, inclusive of difficult-to-reach populations and optimise recovery.

**Methods:** The setting is sub-cities of Addis Ababa and rural districts in south-central Ethiopia covering 1.1 million people and including rural, urban and homeless populations. SCOPE comprises (i) formative work to understand care pathways and community resources (resource mapping); examine family context and communication (ethnography); develop valid measures of family communication and personal recovery; and establish platforms for community engagement and involvement of people with lived experience; (ii) a population-based incidence study, case-control study and cohort study with 12 months follow-up involving 440 people with psychosis (390 rural/Addis Ababa; 50 who are homeless), 390 relatives and 390 controls. We will test hypotheses about incidence rates in rural vs. urban populations and men vs. women; potential aetiological role of khat (a commonly chewed plant with amphetamine-like properties) and traumatic exposures in psychosis; determine profiles of needs at first contact and predictors of outcome; (iii) participatory workshops to develop programme theory and inform co-development of interventions, (iv) evaluation of the impact of early identification strategies on engagement with care (interrupted time series study); (v) a feasibility cluster randomised controlled trial of interventions for people with recent-onset psychosis in rural settings (10-12 clusters; n=80 participants) with 6 months follow-up to inform a future large-scale trial and investigate implementation processes and outcomes, and (vi) two uncontrolled pilot studies to test acceptability, feasibility of co-developed interventions in urban and homeless populations.

## Background

Psychoses, such as schizophrenia, affect more than 23 million people worldwide, contribute substantially to the global burden of disease and are associated with high rates of disability and mortality [1], particularly in low resource settings where most never receive treatment [2]. The onset of psychosis is mostly in late adolescence and early adulthood, increasing the salience to low- and middle-income countries (LMICs) with younger population profiles. Nonetheless, although over 85% of the world’s population lives in Asia, Africa, Latin America, and the Caribbean, less than 10% of psychosis research is carried out in these settings [3]. Consequently, our knowledge of the early stages of psychoses, especially of the basic epidemiology, risk factors, and early course and outcome, is based almost entirely on research from North America, Western Europe, and Australia. Recent evidence indicates that psychoses are highly heterogeneous in their distribution, aetiology, incidence, presentation and outcome [3]. Population-based research in more diverse contexts on incidence, and predictors of onset and outcomes among representative samples of people with psychosis is critical to develop appropriate evidence-based interventions [4].

There has been relative neglect of psychosis in global mental health research [5], with very few methodologically robust population-based studies of psychosis in LMICs [3]. Most existing studies are based either on clinical samples, likely to be highly unrepresentative of the wider population as only a small proportion of people with psychosis access mental health services [6, 7], or on those with long duration of illness in community settings [8]. The ongoing INTREPID II programme in India, Nigeria and Trinidad [9], based on extensive pilot work [10–12], provides methodology that is feasible to implement in diverse LMICs, thus enabling cross-country comparisons. To date INTREPID methods have not been applied in low-income or Eastern African countries. This is an important gap, given the great diversity of health systems, health profiles, burdens of disease, and social, economic and cultural factors across Africa.

In Ethiopia, SCOPE will align with INTREPID epidemiological methods and build on two internationally-recognised community-based studies of people with psychosis: the Butajira course and outcome study (‘Butajira study’) [13, 14] and the PRIME (Programme for Improving Mental health carE) study [15, 16]. In the Butajira study, an epidemiological sample of 359 people with clinician-confirmed diagnoses of schizophrenia was recruited through community case-finding methods and followed up for an average of 10 years [13, 14]. Important findings from this study were: a high treatment gap (90% lifetime), long duration of untreated psychosis (DUP; mean 7.6 years), 7% street homelessness at baseline [8, 17, 18]; low rates of complete remission [13], high suicide attempt rates [19] and higher levels of disability [20, 21], perpetrated violence and violent victimisation [22] and premature mortality (27.7 years of life lost; standardised mortality ratio 302.7) [23] compared with the general population. Transgenerational transmission of disadvantage was shown [24], with high levels of caregiver burden [25], economic impact [26] and experience of stigma [27]. In the PRIME study, 300 people with severe mental illness (85.3% with primary psychosis) were detected in the community [28, 29]. Key findings were lifetime and current access gaps for biomedical care of 41.8% and 59.9%, respectively, with corresponding figures for faith and traditional healing of 15.1% and 45.2% [7]; only 11.3% received minimally adequate biomedical care in current episode [7]; long DUP (median 5 years); high rates of restraint (25.3% in the preceding 12 months); high exposure to traumatic events [7, 30], high lifetime experience of homelessness (36.3%) [7], higher discrimination in urban residents [31]; higher poverty [32, 33] and food insecurity [34] compared to population controls. Both the Butajira and PRIME studies were limited by potential prevalence bias and limited recruitment from religious healing sites.

In the Butajira and PRIME studies, potentially salient psychosocial risk factors and outcomes for the Ethiopian context were not examined. The evidence gaps about potential risk factors (khat use, trauma exposure), targets for intervention (family communication) and outcomes (personal recovery) will now be described.

i. Khat use: Khat leaves contain *Catha Edulis*, an amphetamine-like substance [35]. Khat is widely used across the Horn of Africa and its diaspora [36, 37]. In Ethiopia, use is increasing, with 15.8% nationally reporting current use [38], but over 50% of adults in some districts [39] and high levels in students [40]. In a case-control study from Somalia, age of onset of khat use was associated with clinical ratings of current psychotic symptoms [41], indicating a potential role as a risk factor for onset of primary psychosis. However, the relationship between culturally relevant patterns of khat use [42] and incidence and early course of psychoses has not been investigated.
ii. Trauma: In studies conducted in high-income countries, co-morbidity of post-traumatic stress disorder (PTSD) in people with psychosis ranges from 13-55% [43]. Exposure to traumatic events is a risk factor for developing psychosis [44] and associated with poorer outcomes [45]. In LMICs, traumatic experiences are prevalent among the general population, with an estimated 8% affected by PTSD [46]. In people with psychosis, traumatic experiences are likely to be more common, arising from restraint or coercive treatments [7], accidents or sexual assault [7] or other forms of violent victimisation [22]. The limited evidence available about the role of traumatic exposures in onset and course of psychosis in LMICs indicates variation of the association across setting [47].
iii. Family communication and involvement: High ‘expressed emotions’ from family members, specifically hostility, critical comments and over-involvement, are associated with increased risk of relapse and hospitalisation of people with psychosis [48]. However, a recent meta-analysis [49] identified only one study of expressed emotion from Africa [50]. Important cultural variability of expressed emotion is recognised [49, 51]. Given the crucial role of the family in caregiving in Ethiopia and other African countries [52], local evidence is urgently needed on relevant family communication patterns to inform intervention.
iv. Personal recovery: Defined as *“a way of living a satisfying, hopeful and contributing life even within the limitations caused by illness”*[53], personal recovery is increasingly recognised as an essential goal for intervention [54]. However, the need to explore applicability of this concept in non-Western populations has been noted [55]. A study from Ethiopia explored the concept of recovery in an urban, hospital-based study [56]. However, evidence from a representative sample is required to ensure innovations are based on priorities and values of people with psychosis.

In high-income countries, “early intervention for psychosis” (EIP) models, comprising packages of treatments for people with psychosis at first contact with services, are effective and cost-effective in those settings [57, 58]. There have been a very small number of studies from middle-income countries where intensive specialist-led EIP models were adapted [59–62], but we are not aware of any wider scale-up of these efforts. In any case, these models have been critiqued for not including efforts to achieve earlier interventions, within a ‘critical period’ of 2-3 years from psychosis onset, in order to mitigate the impacts of untreated psychosis [63, 64]. There have been only a small number of evaluations of interventions to reduce DUP in high-income countries, with limited evidence of sustainable impacts [65], except for a high quality study testing a multi-component intervention in Norway [66]. We are not aware of studies from LMIC where impact of interventions on DUP has been evaluated. In LMICs, where DUP is substantially longer [67] and where delayed access to care has been shown to be associated with poorer functional outcomes [68], interventions to reduce DUP assume even greater importance. It has been argued that early intervention models in LMICs should look very different to their high-income country counterparts, focused on a public mental health approach [69].

Efforts to expand access to care in LMICs have included training community members in proactive case identification and linkage with care in rural populations [29] [70], which have the potential to reduce DUP. However, there has been no work on interventions targeting early manifestations of psychosis or application to urban settings or homeless populations. Furthermore, evidence is needed on what interventions are required at first contact with services to optimise outcomes. Our work in Ethiopia has shown that integrating mental health care into primary healthcare in rural settings can expand access to care for people with psychosis [29], that this model is as effective as psychiatric nurse-led care [71] and delivers benefits in terms of functioning, food security [72] reduced suicidality and substantially reduced experience of discrimination and restraint [15]. However, there was minimal impact on symptom severity, 10.6% still reported experience of restraint after one year, and mortality was high. High levels of exposure to traumatic events [30] and undernutrition (23.2% underweight) were also unaddressed [73]. Furthermore, only 30% received ‘minimally adequate treatment’ over 12 months follow-up [15], driven by poverty, the lack of outreach and expectation of cure [74]. We have no evidence of interventions to optimise earlier interventions in urban or homeless populations. In Ethiopia, people who are homeless and have psychosis are trapped in a vicious cycle of poor health due to systematic exclusion from health care [75]. To optimise recovery of people with psychosis in Ethiopia and other LMICs, there is a need to achieve earlier first contact with care and more effective early interventions suited to the sociocultural and economic settings, that address contextually relevant needs for rural, urban and homeless populations.

In SCOPE we aim to pioneer a radical rethinking of early identification and intervention models for people with psychosis at first contact in Ethiopia, grounded in a detailed understanding of contextual needs, inclusive of difficult-to-reach populations, and based on the priorities of people with psychosis.

The specific aims of SCOPE are to:

1. Map community resources, understand help-seeking contexts, and explore concepts of family communication and involvement and personal recovery to develop contextually appropriate measures.
2. Characterise the epidemiology of psychosis in Ethiopia:

a. Determine incidence, needs and presentation of psychosis at first contact;
b. Investigate the role of the urban environment, poverty, khat use, and traumatic experiences during the life course on onset of psychosis;
c. Identify how these exposures and additional factors, including family communication and involvement, impact on personal recovery and outcomes valued by people with psychosis over a 12-month period.
3. Based on evidence from (1) and (2), co-develop contextually grounded interventions for people with psychosis in rural, urban and homeless populations to achieve earlier and better care at first contact;
4. Pilot test interventions:

a. Assess impact of innovative identification strategies on service engagement;
b. In rural populations to determine acceptability, feasibility, affordability, preliminary efficacy estimates and potential scalability and sustainability;
c. In (1) Addis Ababa, and (2) with people who are homeless and have psychosis, to investigate acceptability and feasibility.

## Methods

Ethical approval for SCOPE has been obtained by the Institutional Review Board of the College of Health Sciences, Addis Ababa University (Ref. 001/22/Psy; 12^th^ January 2022) and the Research Ethics Committee of King’s College London (Ref. HR/DP-21/22-26183; 5^th^ April 2022).

The study designs to address the aims include (i) ethnography, resource mapping, piloting and validation studies for novel measures, and theory of change workshops (**Aim 1**); (ii) a population-based incidence study (**Aim 2a),** case-control study (**Aim 2b**) and cohort study with 12 months follow-up (**Aim 2c**) and nested qualitative study (**Aim 2c**), (iii) co-development of contextually-informed innovations **(Aim 3),** (iv) interrupted time series study **(Aim 4a)**; (v) a feasibility cluster randomised controlled trial (RCT) (**Aim 4b**) and (vi) two uncontrolled pilot studies **(Aim 4c)**. See Fig 1.

**Fig 1:**
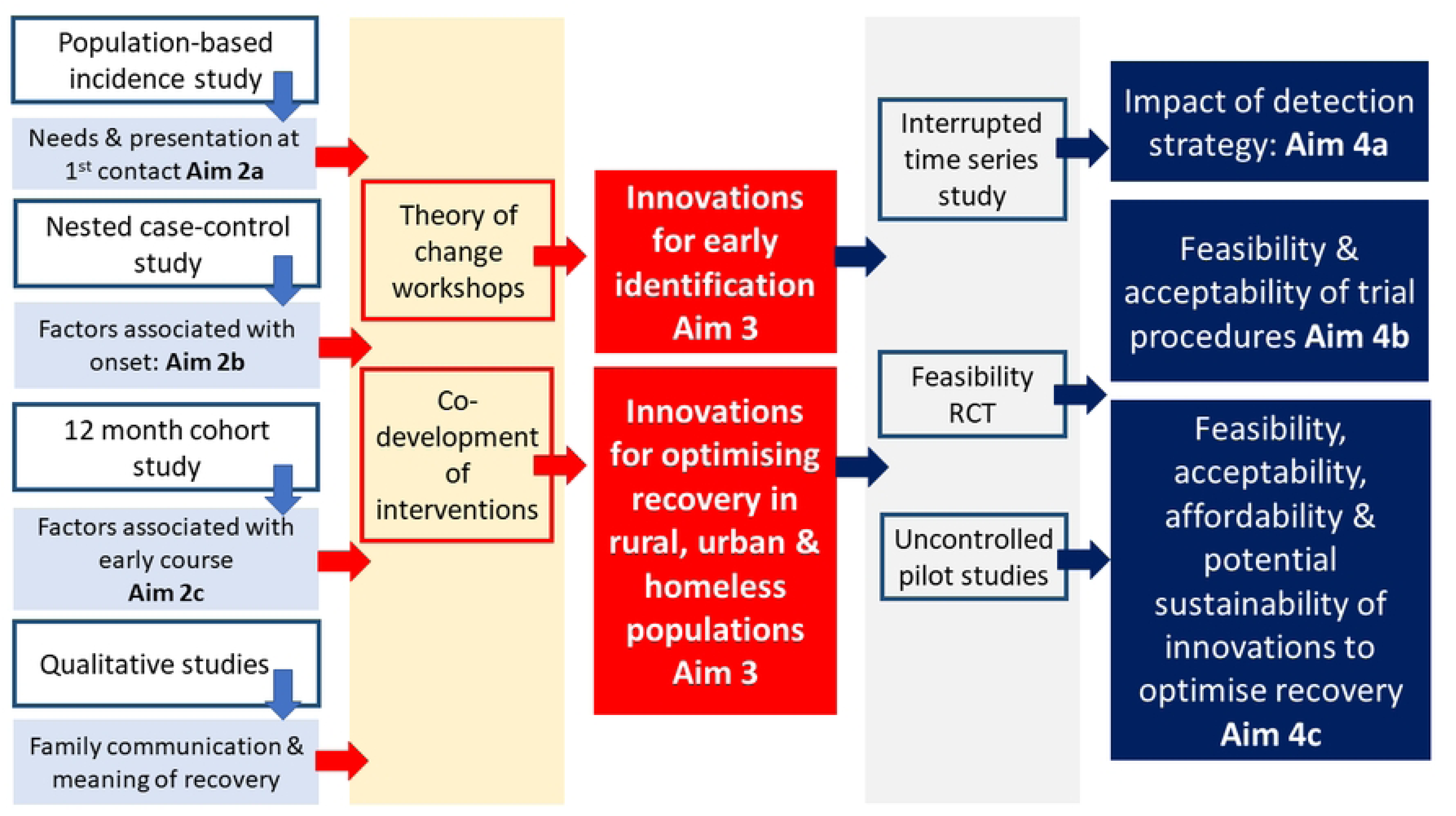
SCOPE study components in relation to aims

## Setting

The studies will be conducted in two sites: (1) contiguous, predominantly rural districts in south-central Ethiopia (Gurage zone of Southern Nations, Nationalities and People’s (SNNP) Region: Misrak Meskan, Merab Meskan, Sodo and South Sodo; Oromia region: Sodo Daci and Kersana Malima; Special district: Sabata Hawas) with an estimated total population of 713,123 people; and (2) Lideta and Kirkos sub-cities of Addis Ababa, the capital of Ethiopia, with an estimated total population of 416,389 in 2016. These settings are described in detail in Supplementary file 1.

## Aim 1: Formative studies and community engagement

The formative work for SCOPE is described briefly below. More detail is provided in Supplementary file 2.

### Resource mapping

Research question: what are the care options, pathways and community resources to support recovery of people with psychosis?

Previous work from Ethiopia identified community resources with potential to be mobilised for people with mental health conditions in rural districts [76]. To map community resources in Addis Ababa, we are using similar methods, including collecting input from the SCOPE community advisory boards, publicly available information, and direct observations. Geographic Information Systems (GPS) coordinates for each potential resource are being recorded using Google Maps [77]

We adapted the PRIME situation analysis tool to document community and health system characteristics across all study districts, complemented by desk reviews, consultations with key informants and the advisory boards [77].

### Ethnographic study

Research question: what are the culturally important aspects of family communication and involvement in relation to people with psychosis in Ethiopia?

Ethnographic observations in households of people with psychosis are combined with in-depth interviews with a range of stakeholders (people with psychosis, caregivers, mental health providers, community leaders). These will investigate patterns of family interaction, impacts of mental ill-health and the status of the individual with psychosis in the family. Field notes from participant observations will be analysed using an interpretative phenomenological approach [78] while interview transcripts will undergo thematic analysis [79] using NVIVO-12 software [80].

We will triangulate findings from both data sources.

### Instrument adaptation and development study

Research question: what is the semantic, content, construct and convergent validity of newly developed measures of (a) family communication and involvement, and (b) personal recovery? For each construct, we will develop, pilot and validate measures using expert consensus meetings, cognitive interviewing, a pilot study and a validation study. Existing measures were first reviewed in light of local qualitative evidence (described above for family communication and involvement, personal recovery evidence based on analysis of data from RISE trial [81]) to produce a list of potentially relevant items. This will be followed by cognitive interviewing with people with psychosis and caregivers to test comprehensibility and acceptability and inform development of a first version of the measures. This will be piloted in 200 people with psychosis and caregivers to test the psychometric properties. After adaptation based on the pilot, the final version will be validated with 400 people with psychosis and 400 caregivers to explore construct validity (using confirmatory factor analysis) and convergent validity with symptom severity and functional impairment. As there is no definitive way to determine sample size for scale validation, depending in part on how well the items are related to the construct under investigation [82], we will follow the recommended rule of thumb to use a participant-to-item ratio of 10:1 [83], assuming that we will have no more than 40 items per scale. Consensus meetings will be held with both Ethiopian and international experts, with academic, clinical, and lived experience, to review study findings at each stage. The resulting measures will be used in the epidemiological study.

#### Community engagement and involvement of people with lived experience

We have established multi-sectoral advisory boards (including people with lived experience, family members, community leaders, traditional and religious healers, police, health care administrators, managers and practitioners, non-governmental sector and non-health government representatives e.g. youth, social care, women’s affairs) in the Oromia and SNNP region rural sites (n=2) and Addis Ababa (n=1) who will provide local oversight and project ownership, support community mobilisation, and enabling trouble-shooting. Project progress and preliminary findings will be presented to the advisory boards on a regular basis to obtain feedback and contribute to the co-production of innovations to be tested in SCOPE.

The SCOPE study has benefitted from input from people with psychosis from the beginning of protocol development. Ongoing involvement is supported through (1) a co-investigator from Ethiopia with lived experience, (2) empowerment activities to support active involvement of people with psychosis from rural districts, (3) supporting the grass-roots service user association in Sodo through enabling meetings and developing peer support, (4) including lived experience feedback as a standing item at advisory board meetings, and (5) close engagement with the Mental Health Service User Association of Ethiopia and Sodo service users in research processes, including planning safeguarding procedures, reviewing information leaflets and case vignettes, instrument adaptation, and co-developing community identification strategies and rights-based interventions.

## Aim 2: Epidemiological study

### Methods for the epidemiological study are closely aligned with the INTREPID study [9, 10]

#### Hypotheses

##### Incidence study

i. Incidence of psychosis will be higher in men [84] and in urban residents [85].

##### Case-control study

(ii) Earlier age of khat initiation and/or problematic khat use [41], exposure to traumatic events prior to psychosis onset and childhood adversity [44] will be associated with increased odds of psychosis

##### Cohort study

(iii) Maladaptive family interactions [49], exposure to traumatic events [45, 86], physical co-morbidity, and poverty will be associated with poorer 12-month clinical, functional and personal recovery, after adjusting for confounders.

#### Sample

We will recruit people with untreated psychosis (rural n=240; urban n=150), relatives (rural n=240; urban n=150) and age- and gender-matched population controls (rural n=240; urban n=150).

Eligibility criteria will be identical to those used in INTREPID [9] with an additional requirement that onset of psychosis was within the past two years, and extending the lower age to 15 years and removing an upper age limit (see Table 1).

**Table 1:** Eligibility criteria for SCOPE epidemiological study.

| Inclusion criteria | Exclusion criteria |
| --- | --- |
| **Cases** |  |
| • Aged 15 years or above<br>• Resident in the catchment area<br>• ICD-11 primary psychotic disorder<br>• Not treated with antipsychotic medication for $\geq 1$ continuous month<br>• Onset within 2 years | • Moderate or severe neurodevelopmental disability (e.g. intellectual disability)<br>• Clinically manifest organic cerebral disorder<br>• Transient psychotic symptoms resulting from acute intoxication |
| **Matched controls** |  |
| • Aged 15 years or above<br>• Resident in the catchment area<br>• Same gender as index case<br>• Age within 5 years of index case | • Past or current psychotic disorder<br>• Moderate or severe learning disability<br>• Clinically manifest organic cerebral disorder |
| **Relatives** |  |
| • Aged 15 years or above<br>• Relative or carer for a person with psychosis recruited into the study | • Insufficient contact with person with psychosis to provide information on family burden and mental health |

In addition, in the Addis Ababa sub-cities we will identify people with untreated psychosis who are homeless. The methodology used for the homeless sample will necessarily differ from the main epidemiological study (as outlined below) but will allow quantification of the proportion of people with untreated psychosis in Addis Ababa who are homeless. On the basis of our previous work, we expect 31 homeless people to have untreated psychosis on initial case-finding (cross-sectionally) with further incident cases recruited during follow-up [87]. See Fig 2 for samples in the different components of the epidemiological study.

**Fig 2:**
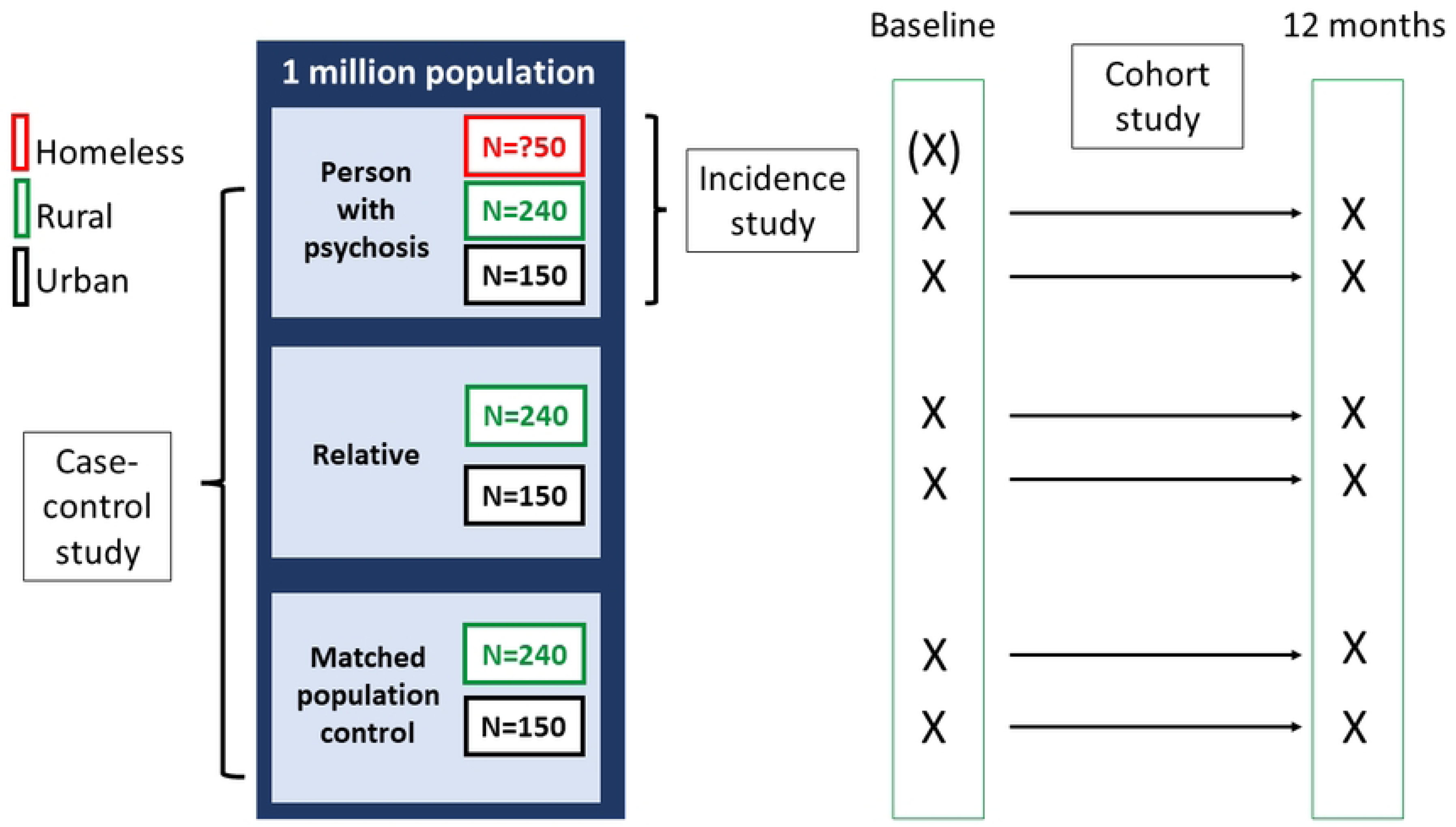
Samples for epidemiological studies

#### Sample size

##### Incidence

With a conservative estimate of the population at risk (≥15 years) of 741,936 (rural population of 713,123 with an estimated 62% aged 15 years or above, and an urban population of 416,389 in 2016 with an estimated 72% aged 15 years or above) screened for incident cases over two years, assuming an incidence of psychosis of 18.7/100000 [84] person-years in the rural setting, this amounts to 139 new cases per year and approximately 554 people with psychosis including those who developed psychosis in the two years preceding baseline. Assuming 70% are identifiable and eligible, we expect to identify around 390 people with psychosis meeting our inclusion criteria. This sample size will have 80% power to detect an incidence rate ratio (IRR) of around 1.6 for urban exposure, in keeping with studies from high-income countries [85], and for males vs. females [84].

##### Factors associated with onset of psychosis (case-control)

For a sample size of 390 people with psychosis and 390 controls, 16% early khat use in controls [38], we will have 80% power to detect an odds ratio of 1.7 for increased odds of early khat use in people with psychosis with p=0.05.

#### Follow-up study

Assuming 20% loss to follow-up, we will have 80% power (p=0.05) to detect a risk difference of 18% in receipt of minimally adequate care (medication prescription and attending for 4 follow-up appointments over 12 months [88]) assuming 30% receipt of minimally adequate care in non-khat chewers [89].

#### Case-finding

Following INTREPID methodology, building on our previous work [76] and arising from community engagement meetings, we will seek to identify people with potential psychosis through a combination of (1) proactive community-based case identification for those not in contact with care, and (2) case identification along help-seeking pathways. Proactive community-based case identification will comprise training of community health extension workers who carry out house-to-house visits for the 1000 households in their geographical catchment area, as well as other community informants, to identify people with possible psychosis. This approach has been shown to be sensitive for identification of people with psychosis in previous work by the Ethiopia team [29, 90]. We will map out places where people with psychosis may seek help, including: health sector (specialist mental health services, general/primary healthcare services, health extension workers, private sector clinics); traditional and faith healing (churches, mosques, holy water sites, other healers); other sectors (police stations, prisons). We will identify focal persons (balanced by gender) in these sites and provide them with orientation and illustrated written materials in local languages. We will ask them to record contact details for any persons with features of psychosis who present to them and to inform the study team. The research team will receive referrals from focal persons and contact them proactively on a monthly basis to identify new presentations of people with possible psychosis. Descriptions of psychosis for both community-based identification and identification on help-seeking pathways will be informed by existing knowledge of local manifestations, extensive stakeholder consultation, including people with lived experience of psychosis, and emerging findings from the epidemiological study [87]. Data collectors will also regularly review health facility contact data supplemented by a registry implemented by the research team to identify people with psychosis.

Our approach to early case identification will be adapted for identification of people who are homeless and have psychosis, mapping out places where people who are homeless commonly congregate, liaising with community police and making contact early in the morning to facilitate privacy [87]. To further facilitate identification, we will also carry out awareness-raising and evidence-based stigma reduction activities in local communities, in collaboration with people with psychosis, and expanding local accessibility of mental health care through integration in primary health care using effective models for this setting [71, 89].

#### Recruitment

##### Person with psychosis and relative

Through linkage with the focal person/key informant, SCOPE data collectors will seek to contact any individuals identified as having probable untreated psychosis with onset in the past two years. For non-homeless persons, the location of households will be identified by working with community-based health extension workers. The person with probable psychosis and a family member will be provided with information about the study. The baseline assessment will then be scheduled. At the baseline assessment, informed consent will be sought. Our approach to assessing capacity and seeking consent builds on previous work [73] and is described below under “key ethical considerations”.

Once recruited, a mental health professional (psychiatrist or master’s level mental health practitioner) will administer a screening questionnaire, including the Psychosis Screening Questionnaire (PSQ), and combine this with clinical observation and relative reports to assess likely eligibility for the main study [91]. The PSQ has been widely used in epidemiological studies to identify persons with probable psychosis. If there is any possibility of psychosis at this stage, the mental health professional will then administer a semi-structured diagnostic interview, the Schedules for Clinical Assessment in Neuropsychiatry (SCAN)[93], to confirm eligibility. A SCAN master trainer from the UK has trained Ethiopian mental health professionals in SCAN. Inter-rater and test-restest reliability will be evaluated. Previous studies have shown that SCAN-trained mental health professionals in Ethiopia have excellent reliability [13].

The full battery of baseline measures (see below) will then be administered to the individual with psychosis and the relative. The assessment will be split over two (or more) sessions to reduce respondent burden.

##### Controls

A control respondent will be identified by mapping the ten nearest neighbouring households and listing all adult residents by gender and age. All potential controls for the case (same sex, age within 5 years) will be approached in random order until an eligible control provides informed consent. This process will be repeated if no consenting control participant is found. The Psychosis Screening Questionnaire will be used to exclude current or past psychotic illness [91].

##### People with psychosis who are homeless

For people who are homeless with probable psychosis, we will find a private place for interview. Consent procedures are described below. People who are homeless and have probable psychosis who can participate in PSQ and SCAN will do so. For those who are too unwell, we will use methodology from our previous study of people who are homeless and have psychosis and base diagnoses on systematic observation using SCAN [87].

All people with psychosis who are identified, regardless of study participation, will be referred to mental health care integrated within local primary health care (PHC) centres or psychiatric care if indicated.

#### Assessment time-points and measures

See Table 2 for measures and assessment time-points. In the PRIME-Ethiopia evaluation of task-shared care for people with psychosis, a 12-month follow-up period was sufficient to model impact of baseline factors on patterns of engagement with care and clinical and social functioning outcomes [7]. Measures denoted with an asterisk* have been adapted and/or validated for the Ethiopian context previously.

**Table 2:** Measures and measurement timepoints.

| Measure | Aim | Baseline |  |  | 12-month follow-up |  |  |
| --- | --- | --- | --- | --- | --- | --- | --- |
|  |  | Case | Relative | Control | Case | Relative | Control |
| Schedules for Assessment in Neuropsychiatry [93]* | 2a | X |  |  |  |  |  |
| MRC socio-demographic schedule | 2a, 2b, 2c | X | X | X | X | X | X |
| Psychiatric and Personal History Schedule [17, 94]* | 2a, 2c | X | X |  | X | X |  |
| WHO Life Chart Schedule [73, 95]* | 2c |  |  |  | X | X |  |
| WHO Disability Assessment Schedule [96, 97]* | 2a, 2c | X | X | X | X | X | X |
| Positive and Negative Symptom Scale (PANSS)[98]* | 2a, 2c | X |  |  | X |  |  |
| Patient Health Questionnaire – 9-item (depressive symptoms) validated for Ethiopia [99] | 2a, 2c | X | X | X | X | X | X |
| Generalised Anxiety Disorder-7 (GAD-7)* [100] | 2a, 2c | X |  |  | X |  |  |
| Global Assessment of Functioning [101]* | 2a, 2c | X |  |  | X |  |  |
| Pre-morbid adjustment scale [102] | 2a, 2b, 2c | X | X |  |  |  |  |
| List of threatening events [103]* | 2b, 2c | X |  | X | X |  | X |
| Alcohol, Smoking and Substance Involvement Screening Tool (ASSIST)[104]* | 2a, 2c | X | X | X | X | X | X |
| Problematic Khat Use Screening Tool* | 2a, 2b, 2c | X | X | X | X | X | X |
| Childhood trauma questionnaire [105] | 2b, 2c | X |  | X |  |  |  |
| Life Events Checklist for DSM-5 and PTSD Checklist for DSM-5 [106] * | 2a, 2c | X |  | X | X |  | X |
| Non-graphic screener for intimate partner violence [107] | 2a, 2c | X |  | X | X |  | X |
| Oslo Social Support Scale*[108] | 2a, 2c | X |  | X | X |  | X |
| Medication checklist [73] |  | X |  | X | X |  | X |
| Glasgow Antipsychotic Side-effect Scale [109, 110] | 2a, 2c | X |  |  | X |  |  |
| WHO-STEPS*[110] | 2a, 2c | X |  | X | X |  | X |
| Family Burden Interview Schedule*[111] | 2a, 2c |  | X |  |  | X |  |
| Laboratory investigation of malaria, anaemia | 2a, 2c | X |  | X | X |  | X |
| Body Mass Index | 2a, 2c | X |  | X | X |  | X |
| Tuberculosis screen | 2a, 2c | X |  | X | X |  | X |
| Adverse events [73] (mortality, homelessness, hospitalisation, victimisation, imprisonment, violence | 2c | X |  |  | X |  | X |
| Household Food Insecurity Access Scale [112]* (HFIAS) | 2a, 2c | X | X | X | X | X | X |
| Camberwell Assessment of Need Short Appraisal Schedule [113]* (CANSAS) | 2a, 2c | X |  | X | X |  | X |
| Discrimination and stigma sub-scale on 'unfair treatment'*[114] (DISC-12) | 2a, 2c | X |  |  | X |  |  |
| NEW: Personal Recovery measure validated for setting | 2a, 2c | X |  |  | X |  |  |
| NEW: Family communication (scale to be selected and adapted) | 2a, 2c | X |  | X | X |  | X |

#### Data collection and management

Mental health professionals trained in SCAN will conduct the diagnostic and semi-structured clinical assessments. Inter-rater reliability will be assessed periodically throughout the study. Lay data collectors with a minimum of completed secondary school education will administer the fully structured instruments. General health workers will collect the laboratory and TB investigations and assess physical health parameters. After training, we will select data collectors who have achieved proficiency. Based on previously developed procedures, we will ensure close supervision and data checking in the field, with weekly data quality checks and queries resolved alongside ongoing data collection. Data collection will be paper based for some aspects of the semi-structured measures, but mostly electronic (using smartphones or tablets) for structured interviews and stored securely on the university server.

#### Data analysis

We will use standard summary statistics, with indicators of spread and precision as appropriate (e.g., crude incidence rates per 100,000 person years, with 95% confidence intervals) to describe the data. We will then use appropriate regression models to compare data between rural and urban settings (e.g., Poisson regression for incidence rates and other count data; Cox regression for time-to-event data; logistic regression (including multinomial) for categorical data (e.g., course type); and linear regression for continuous data (e.g., General Assessment of Functioning score)). In building regression models, we will first fit univariable models, then test for effect modification by pre-specified variables (e.g., gender, age, setting and time) and finally adjust for potential confounders of each hypothesised association by fitting multivariable models.

## Aim 3: Co-development of interventions

The development of innovations to improve early identification and optimise recovery for people with psychosis in rural, urban and homeless populations will follow the Medical Research Council framework for development and evaluation of complex interventions [115]. This process will be informed by: (1) repeated Theory of Change workshops [116] with key stakeholders to harness local knowledge and (2) emerging findings from the epidemiological study, nested qualitative study, and community advisory group meetings, (3) the global evidence base through focused reviews, and (4) co-production workshops.

### Theory of Change (ToC) workshops

We will conduct repeated ToC workshops with key stakeholders in rural and urban sites (each n=25) at months 8, 30 and 59. Stakeholders will include people with psychosis, caregivers, primary healthcare workers and managers, district health administrators, mental health professionals, religious leaders, traditional healers, community representatives, representatives from social welfare, education and law enforcement; non-governmental organisation representatives, and policy-makers. The Ethiopia research team has extensive experience with ToC methodology, including mechanisms to ensure that marginalised voices are heard [117],[118]. Separate ToC roadmaps will be identified for rural, urban and homeless populations. Differing needs for younger and older people with psychosis will be considered. At 30 months, revised ToC maps will identify priority components of contextually relevant models for early identification and intervention for untreated/first-contact psychosis in rural, urban and homeless populations, inform development of a programme theory, and inform the intervention evaluation (process evaluation and outcomes).

Following feasibility testing of interventions, final ToC maps will be produced for each population.

### Co-production workshops

Outputs from the ToC workshop will inform selection of the necessary components of the new interventions and implementation strategies for rural, urban and homeless populations, building on the global evidence base and local experience. People with psychosis will join with small working groups of researchers and practitioners to co-produce interventions, using participatory methods used previously in Ethiopia [119, 120].

## Aim 4a: Interrupted time series study to evaluate identification strategies

We will investigate the impact of SCOPE interventions on identification and linkage of people with untreated psychosis to mental health care using an interrupted time series study design.

We have established recording forms for new cases of psychosis in 65 health facilities across the study site, including information on place of residence, age and gender. We will collect monthly data from the recording forms for the following time periods (1) pre-intervention (7-12 months), during intervention (13-36 months) and post-intervention (37-48 months). We will use segmented regression analysis [121] to examine change in the number of new cases of untreated psychosis contacting mental health care in the study areas, as has been used for previous similar studies. This will allow us to identify changes beyond those related to seasonal effects and secular trends.

## Aim 4b: Feasibility cluster randomised controlled trial

### Research question

How feasible is a randomised controlled trial (RCT) of interventions for people with psychosis at first contact with services; what are implementation processes and outcomes?

### Trial design

Feasibility cluster RCT comparing co-developed interventions for people with untreated psychosis to integrated mental health care within primary care (Enhanced Usual Care: EUC) over a period of six months follow-up, with mixed methods process evaluation.

### Setting

Rural districts in SNNP and Oromia regions.

### Clusters

Clusters will be primary care health centres and their linked catchment areas. There are 23 health centres in the SCOPE rural districts, with variable patient load. We will purposively select 10-12 health centres with higher patient flow, including facilities located rurally and in small towns. The catchment areas of non-selected health centres will be distributed to the selected health centres.

### Sample size

The feasibility RCT will not be powered to evaluate intervention efficacy, but rather to test procedures for a future RCT and delivery of the innovations in routine settings. The impracticality of estimating future parameters for future fully-powered cluster RCTs from pilot/feasibility trials has been noted [122]. From months 37-50 we expect 83 untreated incident cases within the rural districts. We aim to recruit 70-80 participants, giving 6-8 per health centre.

### Intervention group

First contact interventions will be implemented in the selected health centres with linked community health extension workers, as an adjunct to the existing integrated primary mental health care [123],[124]. The first contact interventions will be implemented for six weeks before the trial begins, with PHC workers delivering the interventions to people with psychosis who attend for care but are not eligible for the trial, to embed the intervention, assess provider competence, and ensure fidelity to the model. During the embedding period, weekly group supervision of PHC workers will be convened by a psychiatric nurse. Once the intervention begins, supervision will be monthly, although psychiatric nurses will be available for consultation on complex cases.

### Control group

In the control health centres, PHC workers will be given refresher training using the WHO mental health Gap Action Programme (mhGAP) intervention guideline [124] and receive monthly supervision from a psychiatric nurse (who they can consult about complex cases).

### Recruitment procedures

Following completion of the epidemiological study recruitment, we will continue to detect eligible participants according to the same criteria as the epidemiological study. Their contact details will be recorded using the established SCOPE surveillance system. We will approach both groups in their homes and provide the person and a designated caregiver with information about the RCT. Both will be scheduled to attend a central location for recruitment and baseline assessment if interested. Upon attendance, capacity to consent will be assessed by a mental health professional using established procedures [73]. Informed consent will be obtained, and baseline measures collected. Participants will then be given an appointment at their nearest participating health centre.

### Randomisation

Randomisation of health centres will be stratified by district and location (rural/small town), before participant recruitment starts. An independent statistician will generate the allocation list for sub-districts, which will be stored securely by the trial co-ordinator.

### Masking

It is not possible to mask the study participants or intervention providers to the trial group allocation. Research assessors will be masked to individuals’ group allocation by carrying out assessments in a central location and instructing participants not to say where they live or what type of intervention they have received.

### Participant outcomes

Outcomes for the feasibility RCT will be informed by (1) SCOPE baseline, (2) involvement of persons with lived experience of psychosis, and (3) ToC maps (agreed indicators of success). A key outcome will include engagement in care. Assessments will be carried out at baseline and after 6 months, to allow time for participants to engage with the model to investigate feasibility outcomes.

Analysis of outcomes:

We will use descriptive statistics to present the outcomes in each arm of the trial. Although not the focus of the feasibility RCT, we will use appropriate statistical tests (e.g. t-test, Pearson χ2 test) as exploratory analyses of potential efficacy.

### Process evaluation

A mixed methods process evaluation will be developed based on the ToC model and nested within the trial to investigate (1) feasibility and acceptability of trial procedures, (2) key implementation outcomes: feasibility, acceptability, appropriateness, fidelity, potential scalability and sustainability of the intervention in routine settings, and (3) contextual features of potential relevance to intervention delivery strategies and mechanisms of impact, in light of assumptions identified in the ToC map, based on the Consolidated Framework for Implementation Research (CFIR)[125] (see Table 3).

**Table 3:** Process evaluation measures.

|  | Quantitative | Qualitative |
| --- | --- | --- |
| **Trial procedures** |  |  |
| Numbers screened, eligible, consented, completed baseline | Trial log |  |
| Number who lack capacity | Trial log |  |
| Refusal rate | Trial log |  |
| Acceptability of random allocation | Refusals post-randomisation | X |
| Retention in the trial | Trial log |  |
| Adequacy of masking | Data collector report | X |
| Contamination | Control contact with innovations | X |
| Burden of trial assessments | Duration of assessment | X |
| **Innovations** |  |  |
| Uptake of innovations | % who engage initially |  |
| Retention in treatment | % who remain engaged |  |
| Feasibility, appropriateness and acceptability of innovations in routine settings | Frequency and duration of contacts; Feasibility of Intervention Measure (FIM), Intervention Appropriateness Measure (IAM), Acceptability of Intervention Measure (AIM)[126] | X |
| Fidelity of innovation delivery | Checklists to be developed |  |
| Potential sustainability | Open-ended NOMAD[127] | X |
| **Context** |  |  |
| Contextual mapping & monitoring | Facility profiles; intervention logbooks | X |

### Quantitative data

PHC workers involved in delivering first contact innovations (approximately 40 health centre-based clinicians and approximately 40 health extension workers) will be interviewed using brief structured questionnaires eliciting their views on the intervention acceptability, feasibility and appropriateness [126], and potential sustainability [127].

### Qualitative data

Semi-structured interviews will be conducted with people with psychosis who participated in the trial (control arm, n=5; intervention arm, n=10) and PHC workers (n=8-10), until theoretical saturation is achieved, to explore acceptability and feasibility of trial procedures; and acceptability, feasibility, appropriateness, and potential sustainability and scalability of the intervention in routine settings. Interviews will explore contextual considerations, e.g. in relation to organisational readiness and culture, motivation and opportunity to deliver the intervention, guided by CFIR domains [125] and the ToC map. Interviews will be audiotaped, transcribed in Amharic, and translated into English. Thematic analysis will be used.

### Affordability

We will collect data to estimate intervention costs, including timesheets for intervention staff; detailed utilisation data for participants (Client Service Receipt Inventory^07^); data to calculate unit costs.

## Aim 4c: Uncontrolled pilot studies of interventions for people with psychosis in homeless and urban populations

### Study design

Uncontrolled mixed methods pilot studies to investigate acceptability, feasibility, potential sustainability and perceived impact of innovations for people with psychosis homeless and/or living in urban settings.

### Sample

People with untreated psychosis identified in Lideta sub-city who are 1) homeless or abandoned (n=15) or 2) residing in a household (n= 20).

### Recruitment

We will use the same identification and recruitment procedures as the incidence study over 6 months.

### Intervention

The intervention components developed as an output for Aim 3 will be implemented over 6-8 weeks, focusing on identification, engagement, initial assessment, and acute interventions.

### Assessment

Baseline measures will be the same as for the feasibility RCT and repeated after 6 months.

### Mixed methods process evaluation

We will use the same approach as described for the feasibility RCT, based on the ToC map and using CFIR [125], as indicated in Table 4.

**Table 4:** Process evaluation for pilot studies.

|  | Quantitative | Qualitative |
| --- | --- | --- |
| **Innovations** |  |  |
| Uptake of innovations | % who engage initially |  |
| Retention in treatment | % who remain engaged |  |
| Feasibility, appropriateness and acceptability of innovations in routine settings | Frequency and duration of contacts; Feasibility of Intervention Measure (FIM), Intervention Appropriateness Measure (IAM), Acceptability of Intervention Measure (AIM)[126] | X |
| Potential sustainability | Open-ended NOMAD[127] | X |
| **Context** |  |  |
| Contextual mapping & monitoring | Facility profiles; intervention logbooks | X |

### Quantitative data

Providers of the innovation (approximately 20 community workers and 20 ‘buddies’) will be interviewed using brief structured questionnaires eliciting their views on the intervention acceptability, feasibility and appropriateness [126] and potential sustainability [127].

### Qualitative data

In-depth interviews will be conducted with people who are homeless and have psychosis who participated in the intervention (n=15-20) or providers who contributed to the delivery of the intervention (n=10-15), until theoretical saturation is achieved, to explore the acceptability, feasibility, appropriateness, potential sustainability and scalability of the intervention, and perceived impact. Interviews will be audiotaped, transcribed in Amharic and translated into English. Thematic analysis will be used.

## Key ethical considerations

Mental health services will be established or strengthened in all primary healthcare settings in the study recruitment sites so that persons identified via community case-finding procedures will have access to care locally. To ensure sustainability and integration into the existing health system, we are working closely with community advisory board members, including health administrators, to ensure necessary system supports are in place, including revolving drug funds to access psychotropic medications, training of psychiatric nurses to provide supervision and refresher training to primary healthcare workers, and ensuring that people with lived experience of mental health care are involved in planning and developing services.

Capacity to consent to participate in SCOPE studies will be evaluated by a mental health professional using a systematic and tested approach used previously in our research in Ethiopia [73]. If the individual lacks capacity to consent, a relative will be asked to provide permission for the individual to be included in the study. The justification for this approach is the ethical principle of justice, to ensure that services are informed by the needs of those who are most unwell, balanced against the risk to autonomy which we will minimise by not including people who refuse and re-reviewing capacity. If the person subsequently regains capacity, we will then seek informed consent. If the person refuses participation at that stage, their data will be discarded. We have used this approach previously in Ethiopia [73].

For people who are homeless and have psychosis and who lack capacity to consent, we will identify a designated trusted individual who is well-placed to act in the person’s interests, based on their preference, or, if they are unable to communicate a preference, a professional who is known to them (e.g. community health worker, religious leader). The person with psychosis and designated person will be informed about the study, and the designated person required to give permission.

With our advisory board we will further develop methods for recruiting those who lack capacity and are unknown to community members. We will apply similar safeguards to the persons with psychosis who are not homeless but who lack capacity.

People with psychosis are at increased risk of harms and threats to their human rights in both community and healing settings (biomedical and traditional/faith healing), including restraint, physical or sexual harm, exploitation (financial, sexual, labour), neglect of physical and nutritional health; and unmet basic needs. We have worked closely with the community advisory boards and people with lived experience to develop rights-based safeguarding procedures that are contextually appropriate, to avoid unintended harms.

## Current status

The SCOPE project started formative phase activities on 22^nd^ May 2022 and has completed the ethnographic study, scale development, piloting and validation study, and establishing surveillance methods for early case identification. We anticipate starting the epidemiological study in November 2023.

## Discussion

Findings from the SCOPE project will provide contextually grounded evidence from Ethiopia on the incidence of psychosis, aetiology of psychosis, unmet needs at first presentation and predictors of outcome that could be amenable to intervention. SCOPE will contribute to the small, but growing, number of longitudinal studies of people with psychosis across diverse LMICs. Through use of INTREPID methods, SCOPE data can be used in cross-country comparative analyses to illuminate our understanding of the aetiology and manifestations of psychoses and variations in the types of interventions required. SCOPE will also contribute rich, contextual understanding of the experiences of families affected by psychosis, leading to new measures of family communication and personal recovery that are relevant for this setting. Anchored in strong community engagement and involvement of people with lived experience, evidence generated by SCOPE will feed into development and feasibility testing of interventions (and methods to evaluate interventions) for earlier and better care that optimises recovery for people with psychosis.

The nature of interventions and associated implementation strategies will be informed by emerging data; however, the new interventions for people with psychosis at first contact with services will be located in primary health care rather than specialist mental health services, or sub-specialty early intervention for psychosis services. In part this decision reflects the extremely low population coverage of specialist mental health services in Ethiopia and other LMICs, which contributes to the current inaccessibility of mental health care, long duration of untreated psychosis and high out-of-pocket costs that impoverish families [32]. However, this decision is also informed by the need to promote parity of mental health and physical health care in primary health care and recognition of the benefits to the person with psychosis of staying close to community linkages to address social and economic needs.

Candidate interventions must be rights-based and community-focused, in line with best practice guidance from the World Health Organization [128]. Interventions may draw on critical time intervention (CTI) principles to develop proactive, phased and time-limited approaches to engaging people with psychosis in care to meet the multi-dimensional needs of individuals and their families [129] and/or may draw on elements of community-based rehabilitation with established effectiveness for people with chronic psychosis in rural Ethiopia [130]. Technological innovations have been used to support care for people with psychosis in high-income countries [131] and LMICs [132], although only in specialist settings. mHealth has the potential to link facility-based care with community-based health workers who can provide outreach, information and family support based on our previous materials [123, 133] and facilitate ongoing engagement [132]. Globally, evidence for the contribution of peer support workers is also accumulating [134], and this approach would build on our previous work [117] and ensure relevance and sustainability. Integrated, task-shared approaches to delivering evidence-based brief psychosocial interventions [135–138] also hold promise in LMICs, including Ethiopia, with potential to address traumatic stress symptoms, substance use problems, depressive symptoms, adherence (including family involvement to support adherence [139]) and family support. Innovative decision support tools (technology-guided) and care planning to promote physical health and address under-nutrition may have applicability in mortality reduction.

Interventions for people with psychosis who are homeless will build on previous experience with training community health workers and police for awareness-raising, case-identification and supportive outreach of people who are homeless and have severe mental illness [87]. We will consider innovative ways of addressing unmet physical health, mental health and social needs, while overcoming access barriers. While provision of housing is considered an essential first step to treatment in high-income countries [140], this is not feasible in a low-income country setting, and many do not fulfil criteria for psychiatric admission [87]. Alternative approaches need to be considered to initiate treatment while a person is still homeless. Given the complexity of the SCOPE study, we anticipate that there may need to be modifications made to the protocol to ensure feasibility and achieve the aims. These will be documented, justified and included in publications of the SCOPE findings.

## Conclusions

The SCOPE project seeks to contribute high quality evidence on the sociocultural context of psychosis in Ethiopia and create momentum for earlier and better care through the development and testing of feasible, acceptable and scalable interventions to increase earlier uptake of care and at first contact with services to optimise recovery.

## Data Availability

Deidentified research data will be made available when the study is completed and published.

## Acknowledgments

CH, AA and EM receive support from the National Institute for Health and Care Research (NIHR) through the NIHR Global Health Research Group on Homelessness and Mental Health in Africa (NIHR134325) and CH also receives support from the SPARK project (NIHR200842) using UK aid from the UK Government. The views expressed in this publication are those of the authors and not necessarily those of the NIHR or the Department of Health and Social Care. CH also receives support from WT grant 223615/Z/21/Z. TR receives a fellowship from the British Academy (PF21\210001). CL receives support from the National Institute for Health Research (NIHR) (using the UK’s Official Development Assistance (ODA) Funding) and Wellcome (grant number: 221940/Z/20/Z) under the Department of Health and Social Care (DHSC)-Wellcome Partnership for Global Health Research. CM is part-funded by the ESRC (ESRC Centre for Society and Mental Health at King’s College London: ESRC Reference: ES/S012567/1). For the purposes of open access, the author has applied a Creative Commons Attribution (CC BY) licence to any Accepted Author Manuscript version arising from this submission.

## Data availability

Open access in line with Wellcome policy

## Author contributions

CH, TR, EM, AM, AC, ST, CL, AF, CM, AA conceptualised the overarching research goals and aims, and contributed to funding acquisition. All authors contributed to development of methodology. CH prepared the first draft. All authors contributed to reviewing and editing drafts to produce the final manuscript. All authors approved the final submitted version of the manuscript.

## Supporting Information Captions

Supplementary File 1. Detailed description of settings

Supplementary File 2: Details of formative research

## References

1. McGrath J, Saha S, Chant D, Welham J. Schizophrenia: a concise overview of incidence, prevalence, and mortality. Epidemiologic reviews. 2008;30:67–76. Epub 2008/05/16. doi: 10.1093/epirev/mxn001. PubMed PMID: 18480098.

2. Lora A, Kohn R, Levav I, McBain R, Morris J, Saxena S. Service availability and utilization and treatment gap for schizophrenic disorders: a survey in 50 low-and middle-income countries. Bulletin of the World Health Organization. 2012;90(1):47–54, a-b. Epub 2012/01/25. doi: 10.2471/blt.11.089284. PubMed PMID: 22271964; PubMed Central PMCID: PMCPMC3260570.

3. Jongsma HE, Turner C, Kirkbride JB, Jones PB. International incidence of psychotic disorders, 2002&#x2013;17: a systematic review and meta-analysis. The Lancet Public Health. 2019;4(5):e229–e44. doi: 10.1016/S2468-2667(19)30056-8.

4. Kirkbride JB. Addressing ethnic inequalities in the pathways to care for psychosis. BMC Medicine. 2018;16(1):240. doi: 10.1186/s12916-018-1236-y.

5. Hanlon C. Next steps for meeting the needs of people with severe mental illness in low-and middle-income countries. Epidemiology and Psychiatric Sciences. 2017;26:348–54.

6. Wang PS, Angermeyer M, Borges G, Bruffaerts R, Chiu WT, de Girolamo G, et al. Delay and failure in treatment seeking after first onset of mental health disorders in the World Health Organization’s World Mental Health Survey Initiative. World Psychiatry. 2007;6(3):177–85.

7. Fekadu A, Medhin G, Lund C, DeSilva M, Selamu M, Alem A, et al. The psychosis treatment gap and its consequences in rural Ethiopia. BMC Psychiatry. 2019;19(1):325. doi: 10.1186/s12888-019-2281-6.

8. Kebede D, Alem A, Shibre T, Negash A, Fekadu A, Fekadu D, et al. Onset and clinical course of schizophrenia in Butajira-Ethiopia. A community-based study. Social Psychiatry and Psychiatric Epidemiology. 2003;38:625–31.

9. Roberts T, Gureje O, Thara R, Hutchinson G, Cohen A, Weiss HA, et al. INTREPID II: protocol for a multistudy programme of research on untreated psychosis in India, Nigeria and Trinidad. BMJ Open. 2020;10(6):e039004. doi: 10.1136/bmjopen-2020-039004.

10. Morgan C, John S, Esan O, Hibben M, Patel V, Weiss H, et al. The incidence of psychoses in diverse settings, INTREPID (2): a feasibility study in India, Nigeria, and Trinidad. Psychological Medicine. 2016;46(9):1923–33. Epub 2016/03/28. doi: 10.1017/S0033291716000441.

11. Morgan C, Hibben M, Esan O, John S, Patel V, Weiss HA, et al. Searching for psychosis: INTREPID (1): systems for detecting untreated and first-episode cases of psychosis in diverse settings. Social Psychiatry and Psychiatric Epidemiology. 2015;50(6):879–93. doi: 10.1007/s00127-015-1013-6.

12. Cohen A, Padmavati R, Hibben M, Oyewusi S, John S, Esan O, et al. Concepts of madness in diverse settings: a qualitative study from the INTREPID project. BMC Psychiatry. 2016;16(1):388. doi: 10.1186/s12888-016-1090-4.

13. Shibre T, Medhin G, Alem A, Kebede D, Teferra S, Jacobsson L, et al. Long-term clinical course and outcome of schizophrenia in rural Ethiopia: 10-year follow-up of a population-based cohort. Schizophrenia Research. 2014;161(2-3):414–20.

14. Alem A, Kebede D, Fekadu A, Shibre T, Fekadu D, Beyero T, et al. Clinical course and outcome of schizophrenia in a predominantly treatment-naive cohort in rural Ethiopia. Schizophrenia bulletin. 2009;35(3):646–54. Epub 2008/05/02. doi: 10.1093/schbul/sbn029. PubMed PMID: 18448478; PubMed Central PMCID: PMCPMC2669573.

15. Tirfessa K, Lund C, Medhin G, Hailemichael Y, Habtamu K, Fekadu A, et al. Food insecurity and work impairment in people with severe mental disorders in a rural district of Ethiopia: a cross-sectional survey. Social Psychiatry and Psychiatric Epidemiology. 2019;54(9):1055–66. doi: 10.1007/s00127-019-01709-7.

16. Fekadu A, Hanlon C, Medhin G, Alem A, Selamu M, Giorgis T, et al. Development of a scalable mental healthcare plan for a rural district in Ethiopia. British Journal of Psychiatry Supplement 2015:DOI: 10.1192/bjp.bp.114.153676.

17. Kebede D, Alem A, Shibre T, Negash A, Deyassa N, Beyero T. The sociodemographic correlates of schizophrenia in Butajira, rural Ethiopia. Schizophr Res. 2004;69(2-3):133–41. doi: 10.1016/s0920-9964(03)00089-6. PubMed PMID: 15469186.

18. Kebede D, Alem A, Shibre T, Negash A, Deyassa N, Beyero T. Socio-demographic correlates of bipolar disorder in Butajira, rural Ethiopia. East African medical journal. 2005;82(1):34–9. Epub 2005/08/27. PubMed PMID: 16122110.

19. Shibre T, Hanlon C, Medhin G, Alem A, Kebede D, Teferra S, et al. Suicide and suicide attempts in people with severe mental disorders in Butajira, Ethiopia: 10 year follow-up of a population-based cohort. BMC Psychiatry. 2014;14:150.

20. Kebede D, Alem A, Mitike G, Enquselassie F, Berhane F, Abebe Y, et al. Khat and alcohol use and risky sex behaviour among in-school and out-of-school youth in Ethiopia. BMC Public Health. 2005;5:109. PubMed PMID: 16225665.

21. Kebede D, Fekadu A, Kelkile TS, Medhin G, Hanlon C, Mayston R, et al. The 10-year functional outcome of schizophrenia in Butajira, Ethiopia. Heliyon. 2019;5(3):e01272-e. doi: 10.1016/j.heliyon.2019.e01272. PubMed PMID: 30923757.

22. Tsigebrhan R, Shibre T, Medhin G, Fekadu A, Hanlon C. Violence and violent victimisation in people with severe mental illness in a rural low-income country setting: a comparative cross-sectional community study. Schizophrenia Research. 2014;152:275–82.

23. Fekadu A, Medhin G, Kebede D, Alem A, Cleare AJ, Prince M, et al. Excess mortality in severe mental illness: 10-year population-based cohort study in rural Ethiopia. British Journal of Psychiatry. 2015;206(4):289–96. Epub 2018/01/02. doi: 10.1192/bjp.bp.114.149112.

24. Fekadu W, Craig TKJ, Kebede D, Medhin G, Fekadu A. Multidimensional and intergenerational impact of Severe Mental Disorders. EClinicalMedicine. 2021;41:101151. Epub 20210930. doi: 10.1016/j.eclinm.2021.101151. PubMed PMID: 34632353; PubMed Central PMCID: PMCPMC8488481.

25. Shibre T, Kebede D, Alem A, Negash A, Deyassa N, Fekadu A, et al. Schizophrenia: illness impact on family members in a traditional society - rural Ethiopia. Social Psychiatry and Psychiatric Epidemiology. 2003;38:27–34.

26. Zergaw A, Hailemariam D, Alem A, Kebede D. A longitudinal comparative analysis of economic and family caregiver burden due to bipolar disorder. African Journal of Psychiatry. 2008;11:191–8.

27. Shibre T, Negash A, Kullgren G, Kebede D, Alem A, Fekadu A, et al. Perception of stigma among family members of individuals with schizophrenia and major affective disorders in rural Ethiopia. Social Psychiatry & Psychiatric Epidemiology. 2001;36(6):299–303. PubMed PMID: 11583460.

28. Baron EC, Rathod SD, Hanlon C, Prince M, Fedaku A, Kigozi F, et al. Impact of district mental health care plans on symptom severity and functioning of patients with priority mental health conditions: the Programme for Improving Mental Health Care (PRIME) cohort protocol. BMC Psychiatry. 2018;18(1):61. doi: 10.1186/s12888-018-1642-x.

29. Hailemariam M, Fekadu A, Medhin G, Prince M, Hanlon C. Equitable access to mental healthcare integrated in primary care for people with severe mental disorders in rural Ethiopia: a community-based cross-sectional study International Mental Health Systems. 2020;13:78. 10.1186/s13033-019-0332-5.

30. Ng LC, Medhin G, Hanlon C, Fekadu A. Trauma exposure, depression, suicidal ideation, and alcohol use in people with severe mental disorder in Ethiopia. Social Psychiatry and Psychiatric Epidemiology. 2019;54(7):835–42. doi: 10.1007/s00127-019-01673-2.

31. Forthal S, Fekadu A, Medhin G, Selamu M, Thornicroft G, Hanlon C. Rural vs urban residence and experience of discrimination among people with severe mental illnesses in Ethiopia. BMC Psychiatry. 2019;19(1):340. doi: 10.1186/s12888-019-2345-7.

32. Hailemichael Y, Hailemariam D, Tirfessa K, Docrat S, Alem A, Medhin G, et al. Catastrophic out-of-pocket payments for households of people with severe mental disorder: a comparative study in rural Ethiopia. International Journal of Mental Health Systems. 2019;13(1):39. doi: 10.1186/s13033-019-0294-7.

33. Hailemichael Y, Hanlon C, Tirfessa K, Docrat S, Alem A, Medhin G, et al. Catastrophic health expenditure and impoverishment in households of persons with depression: a cross-sectional, comparative study in rural Ethiopia. BMC Public Health. 2019;19(1):930. doi: 10.1186/s12889-019-7239-6.

34. Tirfessa K, Lund C, Medhin G, Hailemichael Y, Fekadu A, Hanlon C. Food insecurity among people with severe mental disorder in a rural Ethiopian setting: a comparative, population-based study. Epidemiology and Psychiatric Sciences. 2019;28(4):397–407. Epub 2017/11/16. doi: 10.1017/S2045796017000701.

35. Feyissa AM, Kelly JP. A review of the neuropharmacological properties of khat. Progress in neuro-psychopharmacology & biological psychiatry. 2008;32(5):1147–66. Epub 2008/06/20. doi: 10.1016/j.pnpbp.2007.12.033. PubMed PMID: 18561890.

36. Ongeri L, Kirui F, Muniu E, Manduku V, Kirumbi L, Atwoli L, et al. Khat use and psychotic symptoms in a rural Khat growing population in Kenya: a household survey. BMC Psychiatry. 2019;19(1):137. Epub 2019/05/09. doi: 10.1186/s12888-019-2118-3. PubMed PMID: 31064338; PubMed Central PMCID: PMCPMC6505064.

37. Bhui K, Warfa N. Trauma, khat and common psychotic symptoms among Somali immigrants: a quantitative study. Journal of ethnopharmacology. 2010;132(3):549–53. Epub 2010/07/22. doi: 10.1016/j.jep.2010.07.027. PubMed PMID: 20647038.

38. Ethiopia STEPS Survey 2015 Fact Sheet [Internet]. Addis Ababa, Ethiopia: EPHI: http://www.ephi.gov.et/; 2015

39. Alem A, Kebede D, Kullgren G. The prevalence and socio-demographic correlates of khat chewing in Butajira, Ethiopia. Acta psychiatrica Scandinavica Supplementum. 1999;397:84–91. Epub 1999/09/02. PubMed PMID: 10470360.

40. Alemu WG, Zeleke TA, Takele WW, Mekonnen SS. Prevalence and risk factors for khat use among youth students in Ethiopia: systematic review and meta-analysis, 2018. Annals of General Psychiatry. 2020;19(1):16. doi: 10.1186/s12991-020-00265-8.

41. Odenwald M, Neuner F, Schauer M, Elbert T, Catani C, Lingenfelder B, et al. Khat use as risk factor for psychotic disorders: a cross-sectional and case-control study in Somalia. BMC medicine. 2005;3:5. Epub 2005/02/15. doi: 10.1186/1741-7015-3-5. PubMed PMID: 15707502; PubMed Central PMCID: PMCPMC554104.

42. Mihretu A, Teferra S, Fekadu A. What constitutes problematic khat use? An exploratory mixed methods study in Ethiopia. Substance abuse treatment, prevention, and policy. 2017;12(1):17. Epub 2017/03/23. doi: 10.1186/s13011-017-0100-y. PubMed PMID: 28327160; PubMed Central PMCID: PMCPMC5361726.

43. Grubaugh AL, Zinzow HM, Paul L, Egede LE, Frueh BC. Trauma exposure and posttraumatic stress disorder in adults with severe mental illness: a critical review. Clin Psychol Rev. 2011;31(6):883–99. Epub 2011/04/27. doi: 10.1016/j.cpr.2011.04.003. PubMed PMID: 21596012.

44. Varese F, Smeets F, Drukker M, Lieverse R, Lataster T, Viechtbauer W, et al. Childhood adversities increase the risk of psychosis: a meta-analysis of patient-control, prospective-and cross-sectional cohort studies. Schizophrenia bulletin. 2012;38(4):661–71. Epub 2012/03/31. doi: 10.1093/schbul/sbs050. PubMed PMID: 22461484; PubMed Central PMCID: PMCPMC3406538.

45. Mueser KT, Lu W, Rosenberg SD, Wolfe R. The trauma of psychosis: posttraumatic stress disorder and recent onset psychosis. Schizophr Res. 2010;116(2-3):217–27. Epub 2009/11/27. doi: 10.1016/j.schres.2009.10.025. PubMed PMID: 19939633.

46. Ng LC, Stevenson A, Kalapurakkel SS, Hanlon C, Seedat S, Harerimana B, et al. National and regional prevalence of posttraumatic stress disorder in sub-Saharan Africa: A systematic review and meta-analysis. PLOS Medicine. 2020;17(5):e1003090. doi: 10.1371/journal.pmed.1003090.

47. Oloniniyi IO, Weiss HA, John S, Esan O, Hibben M, Patel V, et al. Life events and psychosis: case–control study from India, Nigeria, and Trinidad and Tobago. BJPsych Open. 2022;8(5):e168. Epub 2022/09/16. doi: 10.1192/bjo.2022.562.

48. Butzlaff RL, Hooley JM. Expressed emotion and psychiatric relapse: a metaanalysis. Arch Gen Psychiatry. 1998;55:547–52.

49. O’Driscoll C, Sener SB, Angmark A, Shaikh M. Caregiving processes and expressed emotion in psychosis, a cross-cultural, meta-analytic review. Schizophr Res. 2019;208:8–15. Epub 2019/04/28. doi: 10.1016/j.schres.2019.03.020. PubMed PMID: 31028000.

50. Ewhrudjakpor C. Case studies of family expressed emotion for persons living with schizophrenia in delta state of Nigeria. European Journal of Mental Health. 2009;4(2):247–56. doi: 10.1556/EJMH.4.2009.2.5.

51. Singh SP, Harley K, Suhail K. Cultural specificity of emotional overinvolvement: a systematic review. Schizophrenia bulletin. 2013;39(2):449–63. Epub 2011/12/23. doi: 10.1093/schbul/sbr170. PubMed PMID: 22190078; PubMed Central PMCID: PMCPMC3576159.

52. Mall S, Hailemariam M, Selamu M, Fekadu A, Lund C, Patel V, et al. “Restoring the person’s life’’: a qualitative study to inform development of care for people with severe mental disorders in rural Ethiopia. Epidemiology and Psychiatric Sciences. 2015:DOI: 10.1017/S2045796015001006.

53. Anthony WA. Recovery from mental illness: the guiding vision of the mental health system in the 1990s. Psychosoc Rehabil J. 1993;16:11–23.

54. Slade M, Adams N, O’Hagan M. Recovery: past progress and future challenges. Int Rev Psychiatry. 2012;24:1–4.

55. Gamieldien F, Galvaan R, Myers B, Sorsdahl K. Exploration of recovery of people living with severe mental illness (SMI) in low-income and middle-income countries (LMIC): a scoping review protocol. BMJ Open. 2020;10(2):e032912. Epub 2020/02/06. doi: 10.1136/bmjopen-2019-032912. PubMed PMID: 32019817; PubMed Central PMCID: PMCPMC7044907.

56. Temesgen WA, Chien WT, Valimaki MA, Bressington D. Predictors of subjective recovery from recent-onset psychosis in a developing country: a mixed-methods study. Social psychiatry and psychiatric epidemiology. 2020.

57. Correll CU, Galling B, Pawar A, Krivko A, Bonetto C, Ruggeri M, et al. Comparison of early intervention services vs treatment as usual for early-phase psychosis: a systematic review, meta-analysis, and meta-regression. JAMA psychiatry. 2018;75(6):555–65.

58. Aceituno D, Vera N, Prina AM, McCrone P. Cost-Effectiveness of early intervention in psychosis: systematic review. The British Journal of Psychiatry. 2019;215(1):388–94.

59. Malla A, Iyer SN, Rangaswamy T, Ramachandran P, Mohan G, Taksal A, et al. Comparison of clinical outcomes following 2 years of treatment of first-episode psychosis in urban early intervention services in Canada and India. The British journal of psychiatry : the journal of mental science. 2020:1–7. Epub 2020/07/07. doi: 10.1192/bjp.2020.126. PubMed PMID: 32624012.

60. Guo X, Zhai J, Liu Z, Fang M, Wang B, Wang C, et al. Effect of antipsychotic medication alone vs combined with psychosocial intervention on outcomes of early-stage schizophrenia: a randomized, 1-year study. Archives of general psychiatry. 2010;67(9):895–904.

61. Valencia M, Juarez F, Ortega H. Integrated treatment to achieve functional recovery for first-episode psychosis. Schizophrenia research and treatment. 2012;2012.

62. Brietzke E, Araripe Neto AG, Dias Á, Mansur RB, Bressan RA. Early intervention in psychosis: a map of clinical and research initiatives in Latin America. Brazilian Journal of Psychiatry. 2011;33:s213–s24.

63. Birchwood M, Macmillan F. Early intervention in schizophrenia. The Australian and New Zealand journal of psychiatry. 1993;27(3):374–8. Epub 1993/09/01. doi: 10.3109/00048679309075792. PubMed PMID: 8250779.

64. Malla A, McGorry P. Early Intervention in Psychosis in Young People: A Population and Public Health Perspective. American Journal of Public Health. 2019;109(S3):S181–S4. doi: 10.2105/ajph.2019.305018. PubMed PMID: 31242015.

65. Oliver D, Davies C, Crossland G, Lim S, Gifford G, McGuire P, et al. Can We Reduce the Duration of Untreated Psychosis? A Systematic Review and Meta-Analysis of Controlled Interventional Studies. Schizophrenia bulletin. 2018;44(6):1362–72. doi: 10.1093/schbul/sbx166.

66. Johannessen JO, Larsen TK, McGlashan T, Vaglum P. Early intervention in psychosis: The TIPS project, a multi-centre study in Scandinavia. Psychosis: Psychological approaches and their effectiveness. London, England: Gaskell/Royal College of Psychiatrists; 2000. p. 210–34.

67. Large M, Farooq S, Nielssen O, Slade T. Relationship between gross domestic product and duration of untreated psychosis in low-and middle-income countries. Br J Psychiatry. 2008;193(4):272–8. doi: 10.1192/bjp.bp.107.041863. PubMed PMID: 18827287.

68. Farooq S, Large M, Nielssen O, Waheed W. The relationship between the duration of untreated psychosis and outcome in low-and-middle income countries: a systematic review and meta analysis. Schizophrenia research. 2009;109(1-3):15–23.

69. Farooq S. Early intervention for psychosis in low-and middle-income countries needs a public health approach. Br J Psychiatry. 2013;202(3):168–9. doi: 10.1192/bjp.bp.112.113761. PubMed PMID: 23457178.

70. Jordans MJ, Kohrt BA, Luitel NP, Lund C, Komproe IH. Proactive community case-finding to facilitate treatment seeking for mental disorders, Nepal. Bulletin of the World Health Organization. 2017;95(7):531–6. Epub 2017/04/25. doi: 10.2471/BLT.16.189282. PubMed PMID: 28670018.

71. Hanlon C, Medhin G, Dewey ME, Prince M, Assefa E, Shibre T, et al. Efficacy and cost-effectiveness of task-shared care for people with severe mental disorders in Ethiopia (TaSCS): a single-blind, randomised, controlled, phase 3 non-inferiority trial. Lancet Psychiatry. 2022;9(1):59–71. doi: 10.1016/s2215-0366(21)00384-9. PubMed PMID: 34921796; PubMed Central PMCID: PMCPMC8872807.

72. Tirfessa K, Lund C, Medhin G, Selamu M, Birhane R, Hailemichael Y, et al. Impact of integrated mental healthcare on food insecurity of households of people with severe mental illness in a rural African district: a community-based, controlled before-after study. Tropical Medicine & International Health. 2020;25(4):414–23 10.1111/tmi.13370.. doi: 10.1111/tmi.13370.

73. Hanlon C, Alem A, Medhin G, Shibre T, Ejigu DA, Negussie H, et al. Task sharing for the care of severe mental disorders in a low-income country (TaSCS): study protocol for a randomised, controlled, non-inferiority trial. Trials. 2016;17(1):76. doi: 10.1186/s13063-016-1191-x.

74. Hailemariam M, Fekadu A, Prince M, Hanlon C. Engaging and staying engaged: a phenomenological study of barriers to equitable access to mental healthcare for people with severe mental disorders in a rural African setting. International Journal for Equity in Health. 2017;16(1):156. doi: 10.1186/s12939-017-0657-0.

75. Osei Asibey B, Conroy E, Marjadi B. Health problems and healthcare service utilisation amongst homeless adults in Africa-a scoping review. BMC Public Health. 2020;20(1):594. doi: 10.1186/s12889-020-08648-y.

76. Selamu M, Asher L, Hanlon C, Medhin G, Hailemariam M, Patel V, et al. Beyond the biomedical: community resources for mental health care in rural Ethiopia. PLoS One. 2015;10(5): e0126666.

77. Hanlon C, Luitel NP, Kathree T, Murhar V, Shrivasta S, Medhin G, et al. Challenges and Opportunities for Implementing Integrated Mental Health Care: A District Level Situation Analysis from Five Low-and Middle-Income Countries. PLoS ONE. 2014;9(2):doi:10.1371/journal.pone.0088437.

78. Pietkiewicz IJ, Smith JA, editors. A practical guide to using Interpretative Phenomenological Analysis in qualitative research psychology2014.

79. Braun V, Clarke V. Using thematic analysis in psychology. Qualitative Research in Psychology. 2006;3(2):77–101. doi: 10.1191/1478088706qp063oa.

80. Ltd QIP. NVivo12. In: Ltd QIP, editor.: https://www.qsrinternational.com/nvivo-qualitative-data-analysis-software/home?_ga=2.211866198.333912038.1687859522-955557704.1687859522; 2020.

81. Asher L, De Silva M, Hanlon C, Weiss HA, Birhane R, Ejigu DA, et al. Community-based rehabilitation intervention for people with schizophrenia in Ethiopia (RISE): study protocol for a cluster randomised controlled trial. Trials. 2016;17(1):1–14.

82. MacCallum RC, Widaman KF, Zhang S, Hong S. Sample size in factor analysis. Psychological Methods. 1999;4:84–99.

83. Fabrigar L, Wegener D, MacCallum R, Strahan E. Evaluating the use of exploratory factor analysis in psychological research. Psychological Methods. 1999;4:272–99.

84. Jongsma HE, Turner C, Kirkbride JB, Jones PB. International incidence of psychotic disorders, 2002-2017: a systematic review and meta-analysis. The Lancet Public Health. 2019;4(5):e229–e44. doi: 10.1016/S2468-2667(19)30056-8.

85. Vassos E, Pedersen CB, Murray RM, Collier DA, Lewis CM. Meta-analysis of the association of urbanicity with schizophrenia. Schizophrenia bulletin. 2012;38:1118–23.

86. Mueser KT, Essock SM, Haines M, Wolfe R, Xie H. Posttraumatic stress disorder, supported employment, and outcomes in people with severe mental illness. CNS spectrums. 2004;9(12):913–25. Epub 2004/12/24. doi: 10.1017/s1092852900009779. PubMed PMID: 15616477.

87. Fekadu A, Hanlon C, Gebre-Eyesus E, Agedew M, Haddis S, Teferra S, et al. Burden of mental disorders and unmet needs among street homeless people in Addis Ababa, Ethiopia. BMC medicine. 2014;12(138):20.08.2014.

88. Wang PS, Demler O, Kessler RC. Adequacy of Treatment for Serious Mental Illness in the United States. American Journal of Public Health. 2002;92:92–8.

89. Hanlon C, Medhin G, Selamu M, Birhane R, Dewey M, Tirfessa K, et al. Impact of integrated district level mental health care on clinical and social outcomes of people with severe mental illness in rural Ethiopia: an intervention cohort study.. Epidemiology and Psychiatric Sciences. 2019:10.1017/S2045796019000398.

90. Shibre T, Kebede D, Alem A, Negash A, Kibreab S, Fekadu A, et al. An evaluation of two screening methods to identify cases with schizophrenia and affective disorders in a community survey in rural Ethiopia. International Jounal of Social Psychiatry. 2002;48(3):200–2008. 10.1177/002076402128783244.

91. Bebbington P, Nayani T. The psychosis screening questionnaire. International Journal of Methods in Psychiatric Research. 1995;5(1):11–9.

92. Bitta M, Thungana Y, Kim HH, Denckla CA, Ametaj A, Yared M, et al. Cross-country variations in the reporting of psychotic symptoms among sub-Saharan African adults: A psychometric evaluation of the Psychosis Screening Questionnaire. J Affect Disord. 2022;304:85–92. Epub 20220218. doi: 10.1016/j.jad.2022.02.048. PubMed PMID: 35183621; PubMed Central PMCID: PMCPMC9036658.

93. McGuffin P, Farmer A, Harvey IA. Polydiagnostic application of operational criteria in studies of psychotic illness. Archives of General Psychiatry. 1991;48:764–70.

94. Jablensky A, Sartorius N, Ernberg G, Anker M, Korten A, Cooper JE, et al. Schizophrenia: manifestations, incidence and course in different cultures. A World Health Organization ten-country study. Psychological medicine Monograph supplement. 1992;20:1–97. Epub 1992/01/01. doi: 10.1017/s0264180100000904. PubMed PMID: 1565705.

95. Sartorius N, Janca A, Gulbinat W. Psychiatric assessment instruments developed by the World Health Organization. Social psychiatry and Psychiatric Epidemiology. 1996;31:55–69.

96. Habtamu K, Alem A, Medhin G, Fekadu A, Dewey M, Prince M, et al. Validation of the World Health Organization Disability Assessment Schedule in people with severe mental disorders in rural Ethiopia. Health and Quality of Life Outcomes. 2017;15:64.

97. World Health Organization. WHO psychiatric Disability Assessment Schedule. Geneva: WHO, 1988.

98. Kay SR, Fiszbein A, Opler LA. The positive and negative syndrome scale (PANSS) for schizophrenia. Schizophrenia bulletin. 1987;13:261–76.

99. Kroenke K, Spitzer RL, Williams JB. The PHQ-9: validity of a brief depression severity measure. J Gen Intern Med. 2001;16(9):606–13. doi: 10.1046/j.1525-1497.2001.016009606.x. PubMed PMID: 11556941; PubMed Central PMCID: PMCPMC1495268.

100. Löwe B, Decker O, Müller S, Brähler E, Schellberg D, Herzog W, et al. Validation and standardization of the Generalized Anxiety Disorder Screener (GAD-7) in the general population. Med Care. 2008;46(3):266–74. doi: 10.1097/MLR.0b013e318160d093. PubMed PMID: 18388841.

101. Karterud S, Pedersen G, Loevdahl H. Global assessment of Functioning-Split version (S-GAF): background and scoring manual. Oslo, Norway: Ullevaal University Hospital, Department of Psychiatry,, 1998.

102. Cannon-Spoor HE, Potkin SG, Wyatt RJ. Measurement of premorbid adjustment in chronic schizophrenia. Schizophrenia bulletin. 1982;8:470–84.

103. Brugha T, Bebbington P, Tennant C, Hurry J. The List of Threatening Experiences: a subset of 12 life event categories with considerable long-term contextual threat. Psychological medicine. 1985;15(1):189–94. Epub 2009/07/09. doi: 10.1017/S003329170002105X.

104. Humeniuk R, Ali R, Babor TF, Farrell M, Formigoni ML, Jittiwutikarn J, et al. Validation of the Alcohol, Smoking And Substance Involvement Screening Test (ASSIST). Addiction (Abingdon, England). 2008;103(6):1039–47. Epub 2008/04/01. doi: 10.1111/j.1360-0443.2007.02114.x. PubMed PMID: 18373724.

105. Gossaye Y, Deyassa N, Berhane Y, Ellsberg M, Emmelin M, Ashenafi M, et al. Butajira Rural Health Program: Women’s health and life events study in rural Ethiopia (special issue). Ethiopian Journal of Health Development. 2003;17.

106. Weathers FW, Blake DD, Schnurr PP, Kaloupek DG, Marx BP, Keane TM. The Life Events Checklist for DSM-5 (LEC-5). Instrument available from the National Center for PTSD at www.ptsd.va.gov 2013.

107. Zink T, Levin L, Putnam F, Beckstrom A. Accuracy of Five Domestic Violence Screening Questions With Nongraphic Language. Clinical Pediatrics. 2007;46(2):127–34. doi: 10.1177/0009922806290029. PubMed PMID: 17325085.

108. Dalgard OS, Dowrick C, Lehtinen V, Vazquez-Barquero JL, Casey P, Wilkinson G, et al. Negative life events, social support and gender difference in depression. Social Psychiatry and Psychiatric Epidemiology. 2006;41(6):444–51. doi: 10.1007/s00127-006-0051-5.

109. Waddell L, Taylor M. A new self-rating scale for detecting atypical or second-generation antipsychotic side effects. Journal of Psychopharmacology. 2008;22:238–43.

110. Bonita R, Winkelmann R, Douglas KA, de Courten M. The WHO Stepwise Approach to Surveillance (Steps) of Non-Communicable Disease Risk Factors. In: McQueen DV, Puska P, editors. Global Behavioral Risk Factor Surveillance. Boston, MA: Springer; 2003. p. 9-22.

111. Tessler R, Gamache G. The family burden interview schedule–short form (FBIS/SF). Armherst: Machmer Hall: 1994.

112. Coates J, Swindale A, Blinsky P. Household Food Insecurity Access Scale (HFIAS) for Measurement of Household Food Access: Indicator Guide (v.3). Washington, DC: 2007.

113. Thornicroft G, Beck T, Knapp M, Knudsen HC, Schene A, Tansella M, et al. International Outcome Measures in Mental Health: Quality of Life, Needs, Service Satisfaction, Costs and Impact on Carers. London: Gaskell; 2006.

114. Brohan E, Clement S, Rose D, Sartorius N, Slade M, Thornicroft G. Development and psychometric evaluation of the discrimination and stigma scale (DISC). Psychiatry Research. 2013;208(1):33–40.

115. Skivington K, Matthews L, Simpson SA, Craig P, Baird J, Blazeby JM, et al. A new framework for developing and evaluating complex interventions: update of Medical Research Council guidance. Bmj. 2021;374:n2061. Epub 20210930. doi: 10.1136/bmj.n2061. PubMed PMID: 34593508; PubMed Central PMCID: PMCPMC8482308.

116. De Silva MJ, Breuer E, Lee L, Asher L, Chowdhary N, Lund C, et al. Theory of Change: a theory-driven approach to enhance the Medical Research Council’s framework for complex interventions. Trials. 2014;15(1):267. doi: 10.1186/1745-6215-15-267.

117. Abayneh S, Lempp H, Alem A, Kohrt B, Fekadu A, Hanlon C. Developing a Theory of Change model of service user and caregiver involvement in mental health system strengthening in primary health care in rural Ethiopia. International Journal of Mental Health Systems. 2020;14:51: 10.1186/s13033-020-00383-6.

118. Connell J, Kubisch A. Applying a Theory of Change Approach to the Evaluation of Comprehensive Community Initiatives: Progress, Prospects, and Problems. The Aspen Institute, 1998.

119. Abayneh S, Lempp H, Kohrt BA, Alem A, Hanlon C. Using participatory action research to pilot a model of service user and caregiver involvement in mental health system strengthening in Ethiopian primary healthcare: a case study. International Journal of Mental Health Systems. 2022;16(1):33. doi: 10.1186/s13033-022-00545-8.

120. Abayneh S, Lempp H, Rai S, Girma E, Getachew M, Alem A, et al. Empowerment training to support service user involvement in mental health system strengthening in rural Ethiopia: a mixed-methods pilot study. BMC Health Serv Res. 2022;22(1):880. Epub 20220708. doi: 10.1186/s12913-022-08290-x. PubMed PMID: 35799252; PubMed Central PMCID: PMCPMC9264546.

121. Kokota D, Lund C, Ahrens J, Breuer E, Gilfillan S. Evaluation of mhGAP training for primary healthcare workers in Mulanje, Malawi: a quasi-experimental and time series study. International Journal of Mental Health Systems. 2020;14:3 :10.1186/s13033-020-0337-0.

122. Eldridge SM, Costelloe CE, Kahan BC, Lancaster GA, Kerry SM. How big should the pilot study for my cluster randomised trial be? Statistical Methods in Medical Research. 2016;25(3):1039–56. doi: 10.1177/0962280215588242. PubMed PMID: 26071431.

123. Fekadu A, Hanlon C, Medhin G, Alem A, Selamu M, Giorgis T, et al. Development of a scalable mental healthcare plan for a rural district in Ethiopia. British Journal of Psychiatry Supplement 2016;208(s4–s12):DOI: 10.1192/bjp.bp.114.153676.

124. World Health Organization. Mental Health Gap Action Programme Intervention Guide (mhGAP-IG) for mental, neurological and substance use disorders in non-specialized health settings, version 2.0. Geneva: WHO, 2016.

125. Damschroder LJ, Aron DC, Keith RE, Kirsh SR, Alexander JA, Lowery JC. Fostering implementation of health services research findings into practice: a consolidated framework for advancing implementation science. Implementation Science. 2009;4(1):50. doi: 10.1186/1748-5908-4-50.

126. Weiner BJ, Lewis CC, Stanick C, Powell BJ, Dorsey CN, Clary AS, et al. Psychometric assessment of three newly developed implementation outcome measures. Implementation Science. 2017;12(1):108. doi: 10.1186/s13012-017-0635-3.

127. Finch TL, Girling M, May CR, Mair FS, Murray E, Treweek S, et al. NoMad: Implementation measure based on Normalization Process Theory. [Measurement instrument]. Retrieved from http://www.normalizationprocess.org. Newcastle, UK: Newcastle University, 2015.

128. World Health Organization. Guidance on community mental health services: Promoting person-centred and rights-based approaches. Geneva: WHO, 2021.

129. Herman D, Conover S, Felix A, Nakagawa A, Mills D. Critical Time Intervention: an empirically supported model for preventing homelessness in high risk groups. The journal of primary prevention. 2007;28(3-4):295–312. Epub 2007/06/02. doi: 10.1007/s10935-007-0099-3. PubMed PMID: 17541827.

130. Asher L, Birhane R, Weiss HA, Medhin G, Selamu M, Patel V, et al. Community-based rehabilitation intervention for people with schizophrenia in Ethiopia (RISE): results of a 12-month cluster-randomised controlled trial. Lancet Glob Health. 2022;10(4):e530–e42. doi: 10.1016/s2214-109x(22)00027-4. PubMed PMID: 35303462; PubMed Central PMCID: PMCPMC8938762.

131. Välimäki M, Hätönen H, Lahti M, Kuosmanen L, Adams CE. Information and communication technology in patient education and support for people with schizophrenia. Cochrane Database of Systematic Reviews. 2012;(10). doi: 10.1002/14651858.CD007198.pub2. PubMed PMID: CD007198.

132. Xu D, Xiao S, He H, Caine ED, Gloyd S, Simoni J, et al. Lay health supporters aided by mobile text messaging to improve adherence, symptoms, and functioning among people with schizophrenia in a resource-poor community in rural China (LEAN): A randomized controlled trial. PLOS Medicine. 2019;16(4):e1002785. doi: 10.1371/journal.pmed.1002785.

133. Patel V, Hanlon C. Where there is no psychiatrist. Cambridge, UK: Cambridge University Press; 2018.

134. Puschner B, Repper J, Mahlke C, Nixdorf R, Basangwa D, Nakku J, et al. Using Peer Support in Developing Empowering Mental Health Services (UPSIDES): Background, Rationale and Methodology. Ann Glob Health. 2019;85(1):53. doi: 10.5334/aogh.2435. PubMed PMID: 30951270.

135. Morriss R, Vinjamuri I, Faizal MA, Bolton CA, McCarthy JP. Training to recognise the early signs of recurrence in schizophrenia. Cochrane Database of Systematic Reviews. 2013;(2). doi: 10.1002/14651858.CD005147.pub2. PubMed PMID: CD005147.

136. Pitt V, Lowe D, Hill S, Prictor M, Hetrick SE, Ryan R, et al. Consumer-providers of care for adult clients of statutory mental health services. Cochrane Database of Systematic Reviews. 2013;(3). doi: 10.1002/14651858.CD004807.pub2. PubMed PMID: CD004807.

137. Reilly S, Planner C, Gask L, Hann M, Knowles S, Druss B, et al. Collaborative care approaches for people with severe mental illness. Cochrane Database of Systematic Reviews. 2013;(11). doi: 10.1002/14651858.CD009531.pub2. PubMed PMID: CD009531.

138. Asher L, Patel V, De Silva MJ. Community-based psychosocial interventions for people with schizophrenia in low and middle-income countries: systematic review and meta-analysis. BMC Psychiatry. 2017;17:355: DOI 10.1186/s12888-017-1516-7.

139. Farooq S, Nazar Z, Irfan M, Akhter J, Gul E, Irfan U, et al. Schizophrenia medication adherence in a resource-poor setting: randomised controlled trial of supervised treatment in out-patients for schizophrenia (STOPS). British Journal of Psychiatry. 2011;199(6):467–72. PubMed PMID: 22130748.

140. Hwang SW, Burns T. Health interventions for people who are homeless. The Lancet. 2014;384(9953):1541-7. doi: 10.1016/S0140-6736(14)61133-8.

141. Asher L, Birhanu R, Baheretibeb Y, Fekadu A. “Medical treatments are also part of God’s gift”: Holy water attendants’ perspectives on a collaboration between spiritual and psychiatric treatment for mental illness in Ethiopia. Transcult Psychiatry. 2021;58(4):585–99. Epub 20210525. doi: 10.1177/13634615211015082. PubMed PMID: 34034571.

142. Baheretibeb Y, Soklaridis S, Wondimagegn D, Martimianakis MAT, Law S. Transformative learning in the setting of religious healers: A case study of consultative mental health workshops with religious healers, Ethiopia. Front Psychiatry. 2022;13:897833. Epub 20220913. doi: 10.3389/fpsyt.2022.897833. PubMed PMID: 36177217; PubMed Central PMCID: PMCPMC9513177.

143. ICT Services and System Development and Division of Epidemiology and Global Health. OpenCode 4: Available from: https://www.umu.se/en/department-of-epidemiology-and-global-health/research/open-code2/ (accessed 11th September 2023). 4.03 ed. Umeå, Sweden: Umeå University; 2015.

144. Van Humbeeck G, Van Audenhove C, De Hert M, Pieters G, Storms G. Expressed emotion: a review of assessment instruments. Clin Psychol Rev. 2002;22(3):323–43. Epub 2007/01/05. doi: 10.1016/s0272-7358(01)00098-8. PubMed PMID: 17201189.

145. Burlingame GM, Seaman S, Johnson JE, Whipple J, Richardson E, Rees F, et al. Sensitivity to change of the Brief Psychiatric Rating Scale - Extended (BPRS-E): an item and subscale analysis. Psychological Services. 2006;3(2):77–87.

